# Population Data-Driven Formulation of a COVID-19 Therapeutic

**DOI:** 10.1101/2020.07.24.20161547

**Authors:** Heather R. Campbell, Regan Cecil, Robert A. Lodder

**Affiliations:** Department of Pharmaceutical Sciences, University of Kentucky, Lexington, KY, USA; Department of Biology, University of Kentucky, Lexington, KY, USA

## Abstract

This study is designed to utilize computer modeling of the US population through NHANES to reduce the need for preclinical formulation and toxicology studies of an Ebola anti-viral (BSN389) being repurposed for COVID-19, and to thereby speed the candidate therapeutic to the clinic.

## Introduction

Coronavirus disease of 2019, or COVID-19, is one of six identified coronavirus types to cause disease in humans (Repici et al., 2020). The disease is caused by severe acute respiratory syndrome coronavirus 2 (SARS-CoV-2) and was first identified in patients in Wuhan, China (Repici et al., 2020). Major symptoms include fever, cough, shortness of breath, fatigue, new loss of taste and smell, sore throat, congestion or runny nose, nausea or vomiting as well as diarrhea (Repici et al., 2020; Tina, Saey, 2020a). Over half of patients infected report difficulty breathing, with some reported cases of acute respiratory distress syndrome (Repici et al., 2020; Wu, 2020). COVID-19 is known to be transmitted from person to person through virus-containing droplets that may remain in the air or on surfaces for extended amounts of time (Ghose, 2020; Tina, Saey, 2020b). The exact length of time the virus can survive in the air or on a given surface seems to be highly dependent on the surface itself (Volkin, 2020). Regardless, COVID-19 is thought to be particularly transmittable due to ridged Spike (S) proteins it contains (Saplakoglu, 2020). These S proteins have been found to bind well to the angiotensin-converting enzyme 2 (ACE2) found on the human cell surface, thereby serving as an entry point for the virus (Balfour, 2020; Gray, 2020). The virus can spread asymptomatically and has been predicted to infect millions of people, with some experts predicting up to 70% of the US population will have been infected by August 2021 (Woodward and Miller, Medaris, 2020).

With millions expected to be infected, it is critical to ensure proper testing and diagnosis of COVID-19 to maintain some control of infections. Currently, two major types of tests are being utilized to identify COVID-19, molecular tests and serological tests. The following two sections provide descriptions of each type.

### Molecular testing techniques

Molecular based testing for COVID-19 aims to detect genetic material from SARS-CoV-2. Although different types of molecular testing exist, they all tend to follow the process of first detecting the RNA material, then making copies of the material until an output measurement is produced if the RNA material was present (Kobokovich et al., 2020).

Laboratory Corporation of America was the first of the large national clinical laboratories in the USA to offer a SARS-CoV-2 nucleic acid test (Haskins, 2020). This test utilizes polymerase chain reaction (PCR) technology to detect the virus via nasopharyngeal (NP) or oropharyngeal (OP) aspirates and washes, NP or OP swabs, and bronchoalveolar lavage (Haskins, 2020). RealTime, a different molecular COVID-19 test manufactured by Abbott, also utilizes PCR technology. However, these PCR tests can be time-consuming to perform, so to address this slowness issue, Abbott released ID Now, the first 5-minute test that utilizes isothermal nucleic acid amplification technology to speed up the process (Billingsley, 2020; Koval et al., 2020). However, the accuracy of the ID Now has been questioned (Spaulding, 2020). In addition to these authorized tests, multiple companies such as EverlyWell and OraSure have also developed take-home test/collection kits, with LabCorps Pixel being the first to gain FDA authorization (Hale, 2020; Koval et al., 2020; Spaulding and Mueller, 2020).

### Serological testing techniques

Serological COVID-19 testing aims to identify individuals’ exposure to the virus based on their immune responses (Kobokovich et al., 2020b). Unlike molecular testing, serological testing, specifically antibody testing, has the capability of identifying individuals who were previously infected, which may provide invaluable insight for COVID-19 related tracking and research (Bai, 2020; Billingsley, 2020; Cohut and Godfrey, 2020; Covid-, 2020; Johnson, 2020; Kobokovich et al., 2020b; Wu and Morrow, 2020). Many antibody tests have already obtained emergency use authorization (EUA) from the FDA, such as Cellex’s Cellex qSARS-CoV-2 IgG/IgM rapid test, which was first the antibody test to be authorized (Hale, 2020c). Additional antibody tests that have EUA include Abbott’s Alinity I and Architect SARS-CoV-2 IgG tests, Autobio’s Anti-SARS-CoV-2 rapid test, Bio-Rad Laboratories Platelia SARS-CoV-2 Total Ab, Chembio Diagnostic systems’ DPP Covid-19 IgM/IgG test and more (Colchester and Roland, 2020; FDA, 2020; Fernandez, 2020; Hale, 2020d). The current list of EUAs from FDA is available at https://www.fda.gov/medical-devices/emergency-use-authorizations-medical-devices/coronavirus-disease-2019-covid-19-emergency-use-authorizations-medical-devices.

Numerous research efforts have been initiated to improve serological testing, such as the suggested serological assay for detecting SARS-CoV-2 seroconversion proposed by Amanant and colleagues (Amanat et al., 2020). It should be noted that antibody testing is not meant to diagnose active COVID-19 and can lack specificity, raising questions regarding its reliability (Cairns, 2020; Hale, 2020e; McKinney, 2020; Patel, 2020). These two aspects, along with the difficulty of scaling PCR testing among other factors, prompted the FDA to authorize a new category of testing based on antigens (McKinney, 2020; Patel, 2020). These antigen tests aim to identify the virus by detecting fragments of proteins found on or within the virus, and provide high specificity for COVID-19. However, these tests are not as sensitive as molecular testing methods (McKinney, 2020). Nevertheless, antigen testing could prove to be cheaper and have a higher throughput (Patel, 2020). The first EUA went to Quidel Corporation’s Sofia 2 SARS Antigen FIA (Burton, 2020).

Because of the possibility of false negatives, physicians are told not to rely solely on these COVID-19 test results alone, but also to evaluate their patients and consider diagnostic tests such as CT (computed tomography) scans. CT scans of COVID-19 patients often exhibit some percentage of ground-glass opacity (GGO) patterns (Belfiore et al., 2020; Dai et al., 2020). GGO’s are not, however, unique to COVID-19, and physicians should still take a holistic approach to diagnose COVID-19, i.e., considering patient history or risks of exposure to COVID-19.

Along with properly testing and diagnosing COVID-19, significant effort has been spent on developing a vaccine to combat the virus. The first expected COVID-19 vaccine in China was available for clinical testing by the end of April (Cahill, 2020). Another player in vaccine development is Inovio Pharmaceuticals, which expected to launch clinical trials in April using their INO-4800 vaccine, with results anticipated in September of 2020 (Cahill, 2020). Regeneron is also working diligently to move its clinical trials of REGN3048 and REGN3051 vaccine and treatment up to early summer(Cahill, 2020). Moderna began testing their mRNA-1273 vaccine clinically beginning in April; this vaccine in particular is designed to target the Spike (S) protein of the virus (Cahill, 2020). Additionally, an intranasal vaccine is being developed via animal studies by the biopharmaceutical company Altimmune (Cahill, 2020).

However, vaccines provide no help to those already infected, so significant effort has also been expended on developing pharmaceuticals to combat COVID-19 in those already infected. A novel treatment may take a significant amount of time to develop so many efforts have been focused on drug repurposing. Examples of this include Gilead’s Remdesivir (GS-5734) a general antiviral drug formulated to treat Ebola. This drug treated 761 patients with the virus in a Wuhan hospital (Cairns, 2020) and has performed better than most drugs to date. Another example is Olumiant, a rheumatoid arthritis drug which was identified as a possible treatment of COVID-19 by means of artificial intelligence (Sagonowsky, 2020) . Additionally, Avigan an influenza antiviral drug has seen promising results and has moved into phase III clinical trials (Balfour, 2020b; Lewis, 2020). However, some repurposing drug attempts, like that of the anti-malaria drug hydroxychloroquine, are failing to show efficacy under scientific scrutiny (Allassan, 2020; Hopkins, 2020; Sagonowsky, 2020). Efforts to identify a possible treatment among other experimental drugs are also being conducted. EIDD-2801 is a compound originally developed for treating influenza. EIDD-2801 has shown to interfere with the virus’ reproduction cycle and has shown promising results in the animal testing (Sheahan et al., 2020). A similar story can be told for the compound EDP-1815, which was developed for anti-inflammatory diseases and is in phase 1b trials for psoriasis (Cotrone, 2020; Verbanas, 2020). EDP-1815 has had success with COVID-19 thus far and is in phase 2 trials at the time of writing this paper (Verbanas, 2020). The RECOVERY trial of dexamethasone showed that the drug could reduce COVID-19 patient death risk by one-third in ventilated patients (from 40% to 28%), and by 1/5 in other patients receiving oxygen only, from 25 percent to 20 percent. There was no benefit in the RECOVERY trial to patients who did not need respiratory support (Mahase, 2020).

As these various compounds begin to enter later phase trials, developers must move away from fit-for-purpose (FFP) formulations and move toward development of a final formulation (Marsac, 2019). Formulation is critical in allowing the compound to scale and become a drug product. One method to cut down on formulation development time and cost is to begin with better-informed initial formulations.This paper uses computer population modeling to aid in the development of a smart formulation from the start, one that obviates the need for extensive preclinical formulation and toxicology studies, thereby significantly reducing development time. This study examines the use of such a data-driven approach for a new candidate COVID-19 treatment, BSN389. BSN389 is an FDA designated orphan drug currently in development as an Ebola treatment (Lodder, 2017). The goal of the present study is to determine the current daily exposure of the population to beta cyclodextrin (BCD), and then keep the amount of BCD in the formulation below that level, so drug use does not add significantly to BCD exposure of human subjects.

## Methods

This study is designed to use computer population modeling to reduce the need for extensive preclinical formulation and toxicology studies, and to thereby speed a candidate COVID-19 therapeutic to the clinic. This study updates a previous study with data collected from the latest NHANES (Lodder, 2017). The data compilation and statistical methods used are described in (Cambell, 2020) and summarized below.

### NHANES Data Mining

Data from the Centers for Disease Control (CDC) National Health and Nutrition Examination Survey (NHANES) (ClinicalTrials.gov Identifier: NCT00005154) are combined in this computational experiment with data on shipments from food ingredient manufacturers to evaluate exposure of the US population through foods to a possible pharmaceutical formulation ingredient, beta cyclodextrin (Lodder, 2017). Formulation is indispensable in drug delivery. An inferior formulation can render a drug product inefficacious. A considerable amount of preclinical research must be conducted before a new drug candidate can be evaluated in humans. The cost of these IND-enabling cGxP studies is usually in the range of $3-$5 million. If a misleading drug product formulation is tested preclinically, new iterations of the formulation must be tested again at added cost. BSN389 was devised for treatment of Ebola virus infections. BSN389 is not particularly soluble in water. Therefore, the objective of formulation is to improve the solubility and bioavailability of the active pharmaceutical ingredient (API).

Data-driven computational science can help to lessen the expense of preclinical work. In the absence of extant human exposure data, an array of tests involving acute and chronic toxicology, cardiovascular, central nervous system, and respiratory safety pharmacology must be completed in at least two species before FDA will allow experiments in humans. Nonetheless, for many compounds (such as those originating as natural products) there is a record of human use. In such situations, computer modeling of a population to determine human use may be sufficient to allow phase 0-1 studies with a candidate formulation in humans.

The CDC’s National Health and Nutrition Examination Survey (NHANES) is a group of studies created to assess the health and nutritional levels of adults and children in the United States. NHANES is distinctive in that it incorporates both interviews and dietary information with physical examinations with laboratory results. The NHANES database can be analyzed to determine the distribution of exposures to a food ingredient, and preliminary human formulation experiments conducted at exposures less than those to which the US population is normally exposed through food. These data can be integrated with data extracted from international ingredient shipments to validate the usage model. This report details the data-driven formulation investigation process employing a novel COVID-19 treatment candidate that, unlike vaccines, can be utilized after a patient has been infected with the disease. BSN389’s mechanism of action allows it to be potentially employed against all strains of the virus, an attribute that vaccines might not share.

Consumption data from individual dietary records, detailing food items ingested by each survey participant, were collated by computer in Matlab and used to generate estimates for the intake of BCD by the U.S. population as previously described (Lodder, 2017). A more complete description of the methods appears in

### β-Cyclodextrin Uses in Food and Pharmaceuticals

Cyclodextrins (CDs) can be procured in three common forms: α-cyclodextrin, β-cyclodextrin and γ-cyclodextrin, which are called the first generation (or parent) cyclodextrins. These cyclodextrins comprise six (α), seven (β) and eight (γ) −(1,4)-linked glycosyl units formed into a ring. The ring-shaped molecule is hydrophilic on the outer surface (permitting the cyclodextrin to dissolve in water) and has a nonpolar cavity on the inside, which provides a hydrophobic environment. Cyclodextrins are able to organize inclusion complexes with a diverse array of hydrophobic guest molecules using this hydrophobic cavity. One or two guest molecules can be entrapped by one, two or three molecules of cyclodextrin.

Because β cyclodextrin is used in both food and pharmaceuticals, BCD exposure estimates are more complicated than estimates made just for food. Manufacturers have to be careful when adding what is apparently a minor amount of BCD to a product because that trivial mass may be enough to push a consumer already ingesting BCDs from other sources higher than the Acceptable Daily Intake (ADI) limit.

Every year cyclodextrins are discussed in numerous scientific research articles and meeting abstracts. Many of these reports are about drugs and drug-related products. Table 1 provides a summary of some of the commercially marketed pharmaceuticals formulated with cyclodextrins. In addition, an abundance of novel inventions continue to incorporate cyclodextrins. CDs are well understood from a regulatory point of view, and a monograph for BCD appeared decades ago in both the US Pharmacopoeia/National Formulary and the European Pharmacopoeia (Del Valle, 2004).

**Table 1.** Commercially marketed pharmaceuticals formulated with cyclodextrins throughout the world.

| **Type** | **Drug name** | **Trade name** | **Route of Administration** | **Country** |
| --- | --- | --- | --- | --- |
| **α-Cyclodextrin** | Cefotiam hexetil hydrochloride | Pansporin T | Oral | Japan |
| **β-Cyclodextrin** | Omeprazole | Omebeta | Oral | Europe |
|  | Benexate hydrochloride | Ulgut, Lonmiel | Oral | Japan |
|  | Chlordiazepoxide | Transilium | Oral | Argentina |
|  | Nimesulide | Nimedex | Oral | Europe |
|  | Nicotine | Nicorette | Sublingual | Europe |
| **Sulfobutylether**<br>**β-Cyclodextrin** | Carfilzomib | Kyprolis | Intravenous | Europe,<br>USA |
|  | Ziprasidone<br>Maleate | Geodon,<br>Zeldox | Intramuscular | Europe,<br>USA |
|  | Aripiprazole | Abilify | Intramuscular | USA,<br>Japan |
|  | Voriconazole | Vfend | Intravenous | Europe,<br>USA |
| **2-Hydroxypropyl**<br>**β-Cyclodextrin** | Itraconazole | Sporanox | Oral, Intravenous | Europe,<br>USA |
| **Methylated**<br>**β-Cyclodextrin** | 17β-Estradiol | Aerodiol | Nasal | Europe |
| **2-Hydroxypropyl**<br>**γ-Cyclodextrin** | Diclofenac<br>sodium | Voltaren | Ocular | Europe |

Data collected from (Jambhekar and Breen, 2016; Kishore Shete et al., 2017; Loftsson et al., 2014; Loftsson and Duchêne, 2007; Nerurkar and Naringrekar, 2013)

### Evaluation of β-Cyclodextrin Use

An evaluation of the utilization of BCD by the U.S. population through the approved uses of BCD was conducted. Appraisals of the intake of BCD were calculated using the approved food uses and maximum use level in conjunction with food consumption data included in the National Center for Health Statistics’ (NCHS) 2015-2016 National Health and Nutrition Examination Surveys (NHANES) (USDA, 2012; Bodner-Montville et al, 2006). Estimates of the mean and 90th percentile intakes were obtained for combined representative approved food uses of BCD (see Appendix for lists of food codes used). The intakes were reported for the following population groups:

- infants, age 0 to 1 year
- toddlers, age 1 to 2 years,
- children, ages 2 to 5 years,
- children, ages 6 to 12 years,
- teenagers, ages 13 to 19 years,
- adults, ages 20 years and up,
- total population (all age groups combined, excluding ages 0-2 years),

## Results and Discussion

### Food Usage

The individual food uses for BCD are synopsized in Appendix 1. Food codes representative of each approved use were selected from the Food and Nutrition Database for Dietary Studies (FNDDS) for the appropriate biennial NHANES survey. In FNDDS, the primary (usually generic) description of a given food is assigned a unique 8-digit food code (USDA, 2012) that appears in Appendix 1.

### Food Survey Results

The estimated “all-user” total intakes of BCD from all approved food uses of BCD in the U.S. by population group is summarized in Table 2.

**Table 2.** **Estimated “All-user” Daily Intake (EDI) of BCD in Targeted Foods by Population Group (2015-2016 NHANES Data)**

|  |  |  |  |  |  |  |  |  |
| --- | --- | --- | --- | --- | --- | --- | --- | --- |
| Population Group | N users | N population | % Users | Mean mass (kg) | Mean EDI (g) | 90th % EDI (g) | Mean EDI (g/kg) | 90th % EDI (g/kg) |
| ages 0-1 | 175 | 293 | 59.73 | 11.22 | 0.0019 | 0.0047 | 0.0002 | 0.0004 |
| ages 1-2 | 191 | 291 | 65.64 | 13.59 | 0.0029 | 0.0100 | 0.0002 | 0.0007 |
| ages 2-5 | 555 | 915 | 60.66 | 16.92 | 0.0029 | 0.0093 | 0.0002 | 0.0005 |
| ages 6-12 | 948 | 1505 | 62.99 | 36.58 | 0.0044 | 0.0150 | 0.0001 | 0.0004 |
| ages 13-19 | 768 | 1143 | 67.19 | 67.35 | 0.0080 | 0.0224 | 0.0001 | 0.0003 |
| ages 20 and up | 4169 | 5748 | 72.53 | 79.95 | 0.0153 | 0.0322 | 0.0002 | 0.0004 |
| ages 2 and up | 6440 | 9311 | 69.17 | 66.96 | 0.0124 | 0.0303 | 0.0002 | 0.0005 |

**Table 3.**
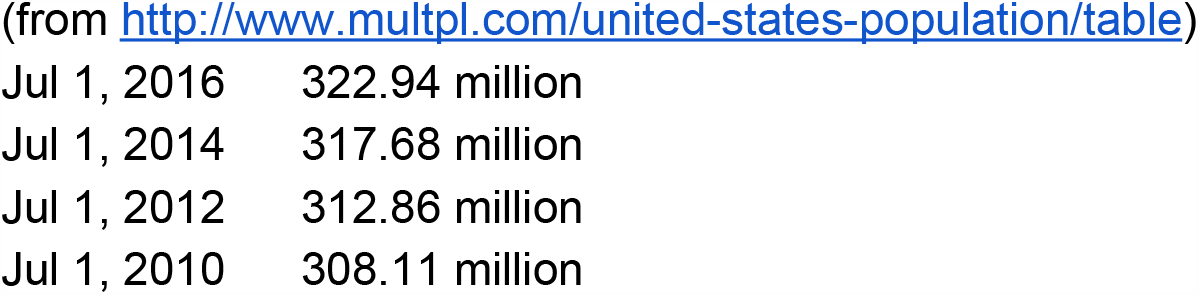
US Population Numbers by Year.

|  |  |
| --- | --- |
| Jul 1, 2016 | 322.94 million |
| Jul 1, 2014 | 317.68 million |
| Jul 1, 2012 | 312.86 million |
| Jul 1, 2010 | 308.11 million |

However, every food category in which BCD is approved for use does not necessarily incorporate BCD into every product in the category at the maximum approved use level. As a result, the values in Figure 1, Figure 2, and Table 2 are corrected using the total amount of BCD consumed in food in the United States during the period of the survey. The correction was derived from US population numbers and market research on the global β cyclodextrin industry (Global and Chinese ν Cyclodextrin Industry, 2016).

**Figure 1.**
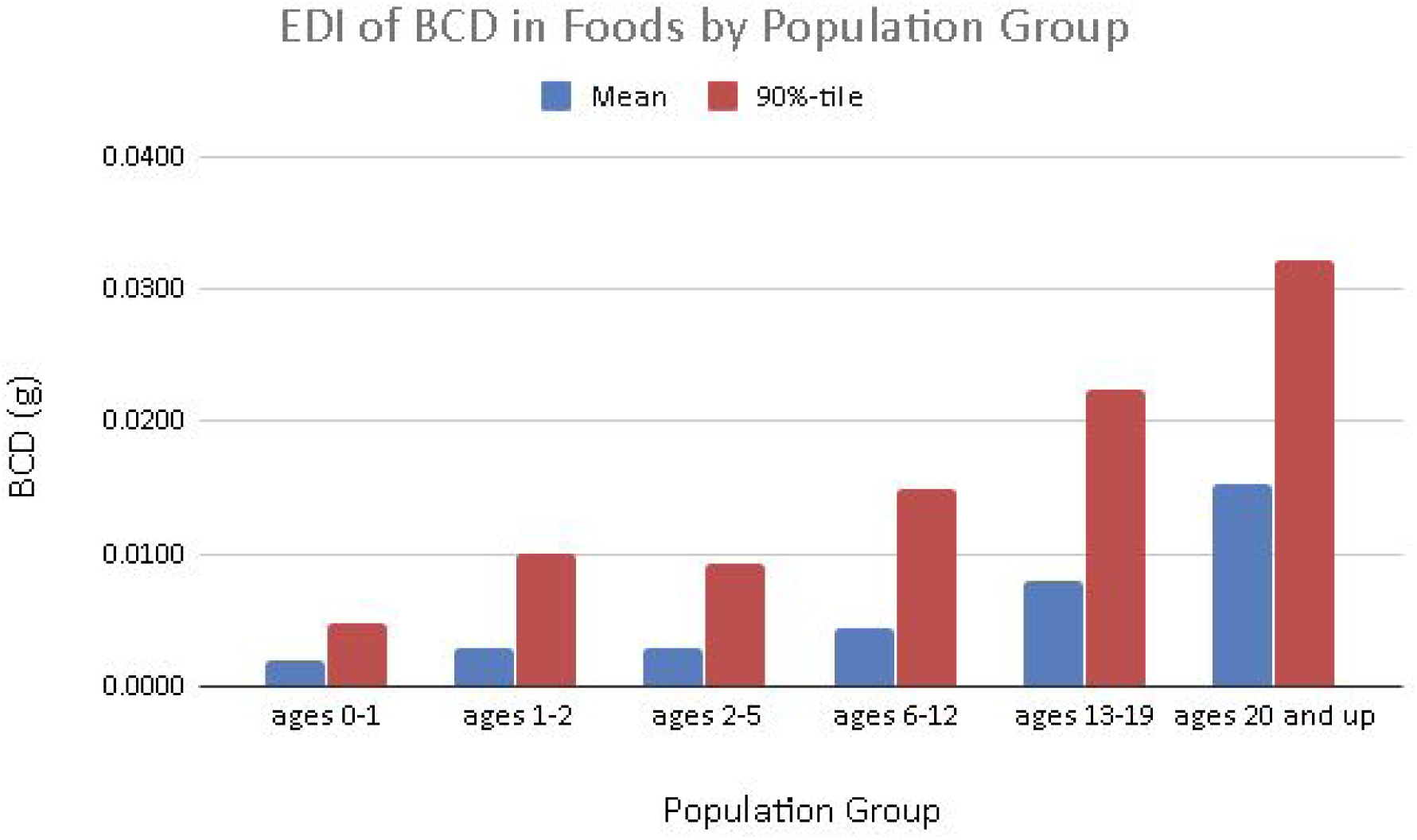
Estimated Daily Intake of BCD in Targeted Foods by Population Group (2015-2016 NHANES Data)

**Figure 2.**
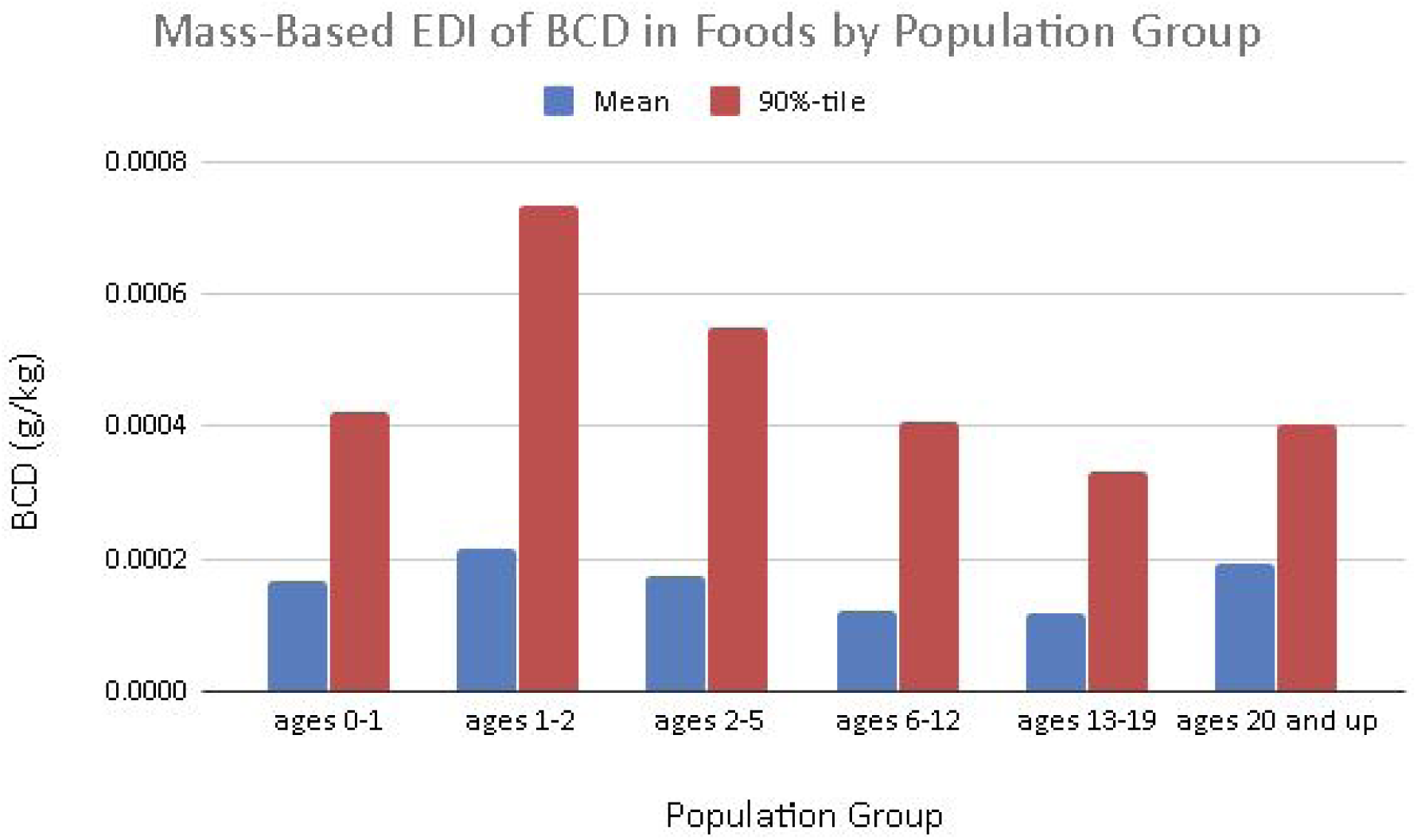
**Weight-Based Estimated Daily Intake of BCD in Targeted Foods by Population Group (2015-2016 NHANES Data)**

(The selected years for the US population numbers correspond to the second year of the biennial NHANES surveys.)

To derive the correction factor the US population number for 2016 was multiplied by the fraction of people aged two and up in the US consuming the targeted food codes:

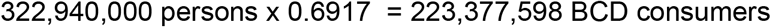

1012 tons of BCD were shipped into the US and presumably consumed in US foods in 2016 (Global and Chinese ν Cyclodextrin Industry, 2016). This number was converted to grams and divided by the number of BCD consumers in the United States to give: 1012000000 g / 223377598 consumers = 4.53 g/consumer/yr.

Dividing the grams per consumer per year by 365 gives grams per consumer per day, or 0.0124 g. To derive the correction factor the maximum g/consumer/day for ages 2+ in the NHANES table is divided by the actual g/day, 2.0237/0.0124 = 163.2. The estimated maximum exposures provided by the 2015-2016 NHANES were divided by 163.2 to get estimated actual exposures in Table 2.

### Development

Scale-up of a novel pharmaceutical may prove difficult during a pandemic, especially with common inactive ingredients such as cyclodextrin, which are used in a wide variety of industries as mentioned (Astray et al., 2009; Hahn, 2020; Loftsson et al., 2014; Loftsson and Duchêne, 2007). *β*-Cyclodextrin, like many active and inactive pharmaceutical ingredients, is now supplied through a global supply chain (GSC) system. Cyclodextrin is made in less than 100 manufacturing sites worldwide, with the majority of these manufacturing sites located overseas in Western Europe and Japan. With a weakened workforce and broken links for importing and exporting goods during the global pandemic, shortages in pharmaceutical products have been reported (Hahn, 2020). Additional research will investigate the feasibility of scale-up of a novel pharmaceutical during periods of material and workforce shortages. This work will construct a model able to identify and mitigate potential supply chain risks and ultimately allow scale-up to occur. The model will need to incorporate country-level reserves and trade data similar to that of the work done by Korniejczuk or Marchand (Korniejczuk, 2019; Marchand et al., 2016). Additionally, the model should capture the actions and interactions of those that will be supplying the material and those that would be further harmed by the shifting inventory adjustments required for producing the new product. This aspect could be feasibly done by integrating social dilemma models such as in Hauser’s work (Hauser et al., 2019).

## Conclusion

BCD use is increasing in terms of the number of foods approved for BCD incorporation, and in 2016 69.17% of the total U.S. population of 2+ years were established as consumers of BCD based on the approved food uses. However, the mean intakes of BCD by the 2016 BCD consumers from all approved food uses were estimated to be 12.4 mg/person/day or 0.2 mg/kg body weight/day, a slight decrease from 2014 (Lodder, 2017). The heavy consumer (90th percentile all-user) intakes of BCD from all approved food-uses were estimated to be 30.3 mg/person/day or 0.5 mg/kg body weight/day. The initial human clinical studies of BSN389 for COVID-19 will use 1.5 μg of BCD. This is over a factor of 1,000 smaller than the expected daily intake from the food uses, and much less than the amount required to move the average consumer from the 50th to the 90th percentile of consumption. For these reasons, the use of BCD in the BSN389 formulation is an negligible inclusion to daily intake and should be safe for subjects in the COVID-19 clinical trial.

## Data Availability

All of the data used were downloaded from the 2015-2016 NHANES (https://wwwn.cdc.gov/nchs/nhanes/ContinuousNhanes/Default.aspx?BeginYear=2015)

## Acknowledgements

The project described was supported by NSF ACI-1053575 allocation number BIO170011 and the National Center for Research Resources and the National Center for Advancing Translational Sciences, National Institutes of Health, through Grant UL1TR001998. The content is solely the responsibility of the authors and does not necessarily represent the official views of the NIH.

